# Perceived Factors Influencing Shared Decision-Making in Mental Health Risk Assessment and Management: A Cross-Sectional Survey with Service Users and Professionals

**DOI:** 10.64898/2026.03.25.26349181

**Authors:** Nafiso Ahmed, Sally Barlow, Lisa Reynolds, Nicholas Drey, Alan Simpson

## Abstract

**Background:** Mental health services are shifting towards person-centred care based on collaboration and shared decision making. Yet evidence indicates that these approaches may not be consistently happening in the assessment and management of risk or safety.

**Methods:** We conducted a cross-sectional online survey to examine perceived barriers and enablers to shared decision-making in risk assessment and management with people living with severe mental illness. Questionnaire development and data analysis were guided by the Theoretical Domains Framework, a psychological framework used to identify and understand factors influencing behaviour change. Items were rated on a 5 int Likert scale. In total, 243 service users and mental health professionals completed the survey.

**Results:** Most service users reported that risk or safety had been discussed with them, but only half felt involved in the risk assessment or management process. Two thirds reported not receiving a copy of their risk assessment or management plan. Service users strongly agreed that communication with professionals about risk and safety requires improvement, and that risk is a difficult and emotive topic to discuss. Professionals reported high motivation to involve service users but identified time constraints and service user related factors as key barriers. Principal component analysis identified four components: (1) motivation; (2) social influences and memory/decision making; (3) beliefs about consequences; and (4) team, environment and training factors. More experienced professionals reported fewer negative beliefs about consequences, such as concerns about causing distress or disengagement.

**Conclusion:** Findings highlight the need for clearer communication, organisational support and targeted training to enhance shared decision-making in risk assessment and management practices.

## Introduction

Risk assessment in mental health care has traditionally focused on identifying potential or imminent risk of harm to self or others including suicidal ideation, self-harm and violence (1-3). Contemporary perspectives, however, have expanded toward a broader understanding of risk that encompasses a wider range of concerns including harassment, stigma, exploitation, systemic or iatrogenic harms, such as restrictive practices or adverse effects of treatment (4). Social risks, including isolation, poverty, housing and discrimination remain a major concern for service users, and underscore the need for a more expansive and context-sensitive approach to risk or safety planning in mental health care (5, 6).

In recent decades, mental health care is shifting from paternalistic models of care towards approaches grounded in recovery, person-centredness and shared decision-making (7). Policy and clinical guidelines call for service users to be actively involved in decisions about their care, including in identifying and managing risk. For example, the UK’s National Institute for Clinical Excellence (NICE) guidelines on self-harm advises against the use of assessment tools and instead recommends a collaborative person-centred approach to understanding and managing risk (8). Building on these principles, NHS England’s Staying Safe from Suicide guidance (9) promotes a holistic, formulation-based approach to safety planning, explicitly advising against risk stratification models and emphasising the importance of understanding each individual’s situation and safety concerns. Similarly, clinical guidance on violence highlights the importance of dynamic, formulation-based risk assessment that incorporates service user perspectives and prioritises clear communication (1). Internationally, collaborative safety planning is considered best practice in both USA and Australia (10, 11).

Nonetheless, research suggests that service users are often not involved in identifying safety concerns, or aware of the information included in their risk management plan (4, 12, 13). A lack of involvement limits the potential benefits of collaborative approaches such as reduced suicidal ideation, strengthening of therapeutic alliance, and enabling more personalised risk formulation (14, 15). It also constrains the identification of broader risk concerns from multiple perspectives, including social and systemic harms.

Shared decision-making in risk assessment and management is widely endorsed by both professionals and service users; however, qualitative research shows its implementation is hindered by a range of contextual, relational, and systemic factors (16, 17). Quantitative research systematically examining these barriers and enablers remains scarce. Therefore, the current cross-sectional survey, guided by the Theoretical Domains Framework (TDF) (18, 19), investigates the factors that influence shared decision-making in risk assessment and management.

This study addresses the following research questions:

1. What are mental health professionals’ and service users’ perceptions of shared decision making in risk assessment and management?
2. What do mental health professionals and service users perceive to be barriers and enablers to shared decision-making in risk assessment and management with people living with severe mental illness?
3. What underlying factors influence mental health professionals’ implementation of shared decision-making in risk assessment and management, and do these factors differ by profession, age, or level of experience?

## Methods

### Design

An exploratory, cross-sectional, online survey was developed and distributed using Qualtrics software (Provo, UT). Data collection and analysis was guided by the Theoretical Domains Framework (TDF) (18, 19), as detailed in our earlier work (13, 17). This survey constitutes the second phase of a sequential mixed methods design (20). Ethical approval to conduct the study was granted by City, University of London’s School of Health Sciences Research Ethics Committee (ETH1920-0144). The study is reported in accordance with the Checklist for Reporting Results of Internet E-Surveys (CHERRIES) (21).

### Participants

People were eligible to take part in interview if they were a mental professional or service user who either worked within or received care and treatment from a community mental health team. Service user participants were eligible if they were aged 18 years or over and living with a severe mental illness defined as psychosis, schizophrenia, bipolar disorder, or major depression. Psychiatrists, psychologists, nurses, social workers, or occupational therapists, whose role involved assessing and managing risk with individuals with severe mental illness were eligible to participate. Professionals were also asked whether they acted as care-coordinators, a role typically undertaken by mental health nurses or social workers who coordinate the care provided by the multidisciplinary team and other providers. The full eligibility criteria are outlined in Table 1.

[INSERT Table 1. Eligibility criteria for cross-sectional survey]

### Data measurements

The survey included items on (1) socio-demographic characteristics and (2) perceived barriers and enablers to shared decision-making in risk assessment and management.

#### Sociodemographic data

From both groups, we collected sociodemographic characteristic data including age, gender and ethnicity. Mental health professional participants were also asked for information about their role, country of practice, type of community mental health service they worked within, experience-related variables (e.g., time qualified, current role duration, care coordinator status), involvement in risk assessment and management, proportion of caseload with severe mental illness (SMI), and training received. Service user participants provided their diagnosis, clinical characteristics (e.g., duration of current support, length of contact with mental health services), and contextual factors such as living situation, relationship status, and frequency of contact with carers. A full summary is provided in Table 2, and 3.

#### Development of the Theoretical Domain Framework survey

Questionnaire development was informed by belief statements from our previous qualitative interview studies (13, 17) and the Theoretical Domains Framework (TDF) (18). The TDF is a synthesis of 33 theories of behaviour change comprising 14 theoretical domains found to influence behaviour. Items were selected through a consensus approach following three criteria: (1) relatively high frequency of specific beliefs; (2) presence of conflicting beliefs; and (3) evidence of strong beliefs that may affect the target behaviour (22).

A 39-item questionnaire was developed for mental health professionals, covering 13 of the 14 TDF domains previously identified as relevant in the interview study (see Additional file 1, Appendix A). A single belief statement related to behavioural regulation, mentioned by one participant in the prior interview study was not included as an item in the survey. Survey items were scored on a 5-point Likert scale from 1 (strongly disagree) to 5 (strongly agree). Negatively worded items were reversed-scored.

Service user respondents were directed to one of two surveys depending on their responses to four initial screening questions: (1) Has anyone ever discussed risk with you? (2) Has anyone ever carried out a risk assessment with you? (3) Has anyone involved you in planning how to manage risk or safety? and (4) Have you ever been given a copy of your risk assessment and risk management plan?

Respondents who answered ‘no’ or ‘not sure’ to all four were presented with a 21-item survey exploring hypothetical beliefs. Those who answered ‘yes’ to any of the questions received a 20-item version based on their lived experience (see Additional file 1, Appendix B for the service user survey). Items were also scored on a 5-point Likert scale and negatively worded items were reversed-scored.

#### Piloting

The questionnaires were piloted on four academics and four members of the target population (i.e., two qualified mental health professionals and two service users). Each reviewed the survey and provided feedback in relation to the relevance and appropriateness of the questions, as well as the technical functionality of the survey. Based on this feedback, improvements and refinements were made to the survey prior to data collection. Detailed feedback from piloting and changes made provided in Additional file 1, Appendix C.

### Procedures

A convenience sampling method was used to recruit eligible participants online via an anonymised survey link. Two charities associated with the service user participant group (i.e., the National Survivor User Network and MQ Mental Health) shared the survey link through their website and electronic newsletter. Additionally, the link was posted on social media platforms including Facebook and X (formerly Twitter), where it was circulated by relevant mental health organisations and academics. The survey link was shared within closed support groups on Facebook for either individuals living with SMI or mental health professionals such as nurses, social workers, psychiatrists or occupational therapists. The survey was open from August 2019 to December 2019. Participants who completed the survey were invited to enter a prize draw to win one of two £25 Amazon vouchers.

### Data analysis

#### Internal consistency of the TDF survey

The internal consistency for each TDF domain was assessed using Cronbach’s alpha (acceptable values ≥0.7) (23). Cronbach alpha is only appropriate for domains that include more than two question items and may yield lower values for scales with fewer than ten items. Therefore, mean inter-item correlation was also calculated where alpha values were <0.7, aiming for an optimal range between 0.15 - 0.5 (Clark & Watson, 2016).

#### Descriptive statistics

Analysis was conducted in IBM SPSS Statistics 26 (24). Descriptive statistics (i.e., percentages, ranges, frequencies, and means) summarised participant demographics and responses. Mean scores for each Likert question were calculated by summing all scores for all items within the domain (strongly disagree = 1 to strongly agree = 5) and divided by the total number of responses. A higher mean score indicated greater agreement with the statement (25, 26). An overall mean score was calculated for each of the domains; negatively worded items were reverse scored.

#### Inferential statistics

Principal Components Analysis (PCA) with Oblimin rotation was used to reduce a large number of variables to a smaller number of components and to test the validity of the questionnaire i.e., whether the principal components support the predefined structure of the TDF (or the broader COM-B model) which underpin the research (18). The COM B model conceptualises behaviour as arising from interactions between Capability, Opportunity and Motivation, providing a higher level behavioural framework against which the derived components were interpreted (27, 28).

The number of components to be retained was decided based on the Kaiser criterion (Eigenvalues >1), visual inspection of the scree plot and meaningfulness of the results according to the theoretical framework. The Kaiser-Meyer-Olkin (KMO) statistics and Bartlett’s Test of Sphericity were used to assess the suitability of the data for PCA (23). The analysis included items that were not freestanding, cross-loading or decreasing the scale’s internal reliability, and that displayed acceptable communalities. Comrey and Lee (29) suggest that factor loadings in excess of 0.71 are excellent, 0.63 are very good, 0.55 are good, 0.45 are fair, and 0.32 are poor. Tabachnick and Fidell (30) recommend that researchers consider replicability, utility, and complexity in selecting factors for interpretation. Therefore, items with factor pattern/coefficients of ≥0.6 were selected, and the TDF/COM-B components guided the selection of items. Full PCA procedures, factor loadings, scree plot and reliability values are provided in Additional file 1, Appendix D.

#### Group comparative analysis of influencing factors

Previous research indicates that some professionals do not perceive it as part of their role to discuss suicidality with service users (31, 32), or risk focus can differ between professionals due to age and experience (3). Therefore, independent t-tests and one-way ANOVAs were used to examine differences in component scores across demographic variables (e.g., age, profession, role, years qualified and years in role). P-values ≤0.05 were considered statistically significant, which is the most commonly used threshold to differentiate between significant and non-significant results (33).

Total component scores were calculated by adding scores from all the items that made up a component (reverse scoring negative items). In order to have sufficient numbers to test for differences in component scores, some of the continuous variables were collapsed into equal groups. The visual binning function in SPSS was used to identify suitable cut-off points to break the continuous variables into three approximately equal groups. Age was categorised into three groups (≤36, 37-46 and 47+), years qualified (≤6, 7-15 and 16+) and years in current role (≤1, 2-4 and 5+). Participants were also grouped as either (1) mental health nurses, (2) social workers and (3) other, including occupational therapists, psychologists, psychiatrists and other. An even number of participants reported being care-coordinators and non-care-coordinator. Subgroup analysis by demographic variables was undertaken to ascertain the presence of any variation in the component scores.

## Results

### Participation

For service user respondents, seventy-two people visited the first page of the survey, of whom 59 (81.9%) consented into the study. Seven people who had consented were excluded as they reported not receiving treatment from an adult community mental health service. Eleven participants had partially completed their questionnaires, of whom seven answered more than the demographic questions. Therefore, data analysis was performed on 48 (66.6%) service user questionnaires.

Three hundred and sixty-three people visited the mental health professional survey’s welcome page, and 343 consented into the study (94%). Fifty-three people were subsequently excluded as they reported not working in an adult community mental health service (n=41), not providing care for service users with SMI (n=9) or that their role did not involve assessing and managing risk (n=3). Ninety-five (28% of 343) participants had partially completed their questionnaire, of whom 20 (6%) completed 50% of the questionnaire. Data analysis was performed on the 195 (57%) completed questionnaires, as this was above the number (n = 159) required to achieve 80% power for the study and is also more than the minimum requirement for principal component analysis (n = >150).

### Characteristics of the respondents

The majority of service user respondents (n=48) were female (n = 39, 81.3%), of white British ethnicity (n = 40, 83.3%) and between the age of 20-58 (M:38, SD:10.7). Bipolar disorder was most frequently reported (n = 16, 23.5%), followed by ‘depression with psychotic features’ (n = 14, 20.6%). Thirty-three (68.8%) service user respondents reported that risk or safety had been discussed with them, most commonly by their care-coordinator (n = 26, 39.4%), followed by a psychiatrist (n = 21, 31.8%). Half (n = 24, 50%) reported being involved in the risk assessment process, again primarily with a care-coordinator (n = 18, 42.9%) or psychiatrist (n=14, 33.3%). Nearly half (n= 23, 47.9%) reported being involved in managing risk/safety. However, two-thirds (n = 32, 66.7%) reported not being given a copy of their risk assessment and management plan. Further characteristics of service user participant are presented in Table 2, and Additional file 1.

Table 3 shows the characteristics of participating mental health professionals. 173 (88.7%) were female and of white, British ethnicity (n=157, 80.5%), and reported primarily practicing as social workers (n=71, 36.4%) or mental health nurses (n=62, 31.8%) in England (77.9%). Most reported having been qualified for more than ten years (n=103, 52.8%), and half of the sample said they were care-coordinators (n=98, 50.3%).

### Results for service users perceived barriers and enablers

#### Internal Consistency of Domains

For the hypothetical questionnaire, Cronbach alpha values for the domain measures ranged from 0.33 (emotions) to 0.87 (beliefs about consequences). For the lived-experience questionnaire, only four domains consisted of more than two items and all their alpha values were marginally < 0.7 (knowledge = 0.68, reinforcement = 0.69, goals = 0.61, social influences = 0.60). Mean inter-item correlations were within the optimal range for nearly all the domains.

#### Questions based on the hypothetical belief statements

Figure 1 illustrates responses to the hypothetical belief statements; mean scores are summarised below to highlight overall patterns of agreement and disagreement. Mean values were mostly above 4 (strongly disagree = 1 to strongly agree = 5) (Table 4), indicating generally favourable views towards being involved in the RA and RM process. The item *"we need better communication between service users and professionals about risk"* (social influences) received an absolute rating of ‘strongly agree’. The first three items measuring service user’s awareness (knowledge) of the content of their RA (M: 4.58, SD: 0.52), the content of their RM (M: 4.75, SD: 0.45) and whether they had received a copy of the documents (M: 4.92, SD: 0.29) received a high mean value.

**Figure 1.**
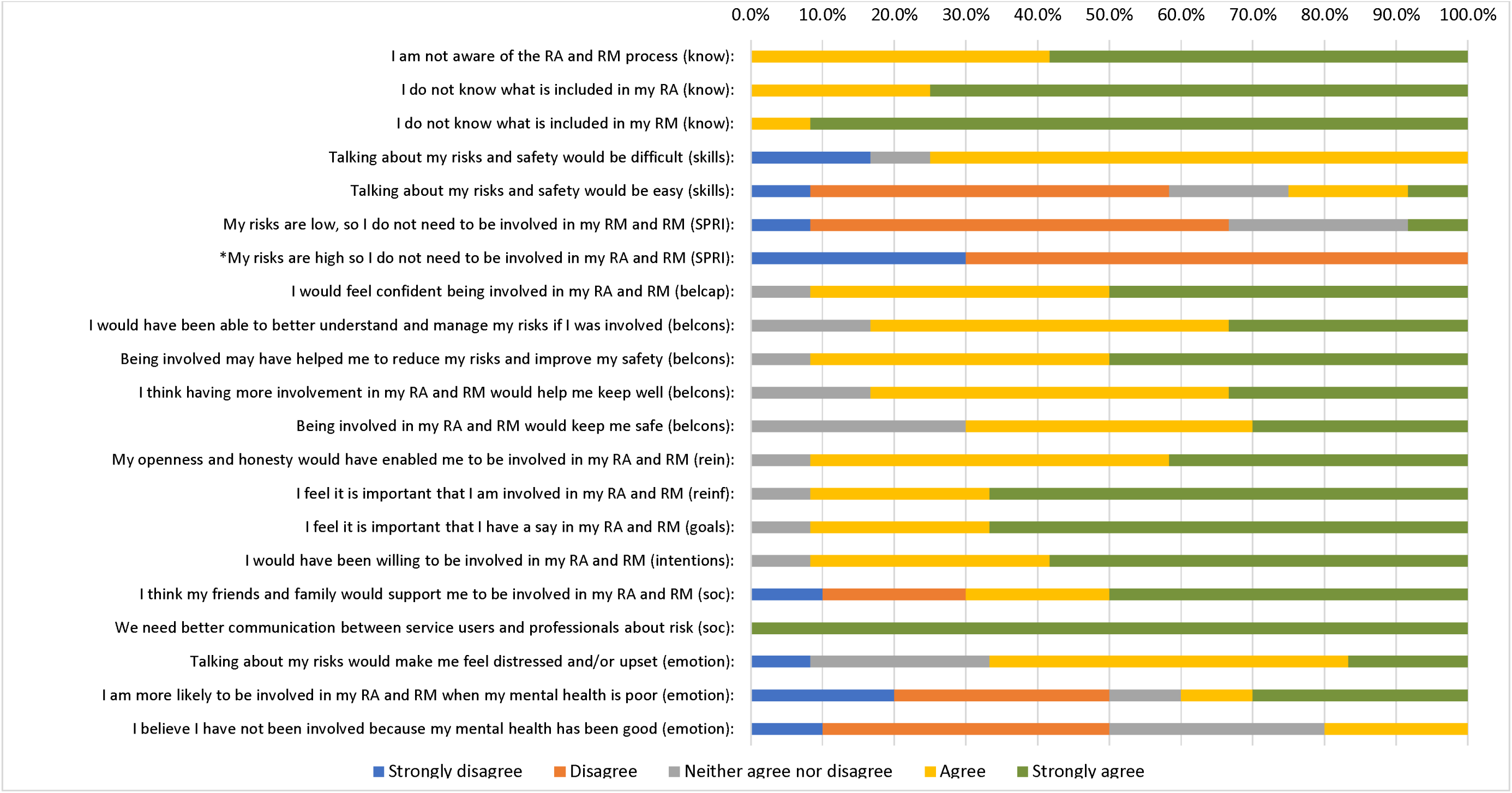
Responses to questions based on the hypothetical belief statements (n= 12)

These items were negatively worded, so the high mean value indicates a lack of knowledge and a potential barrier to involvement. Other items that had high agreement were related to goals *"I feel it is important that I have a say in my risk assessment and risk management"* (M: 4.58, SD: 0.67), and reinforcement *"I feel it is important that I am involved in my risk assessment and risk management"* (M: 4.58, SD: 0.67). The item with the lowest mean value was *"my risks are high, so I do not need to be involved in my risk assessment and risk management"* (M: 1.70, SD: 0.48).

[INSERT Table 4. Mean and standard deviation for each item (hypothetical)]

#### Questions based on lived experience belief statements

Responses to the lived-experience questionnaire are shown in Figure 2, with mean scores summarised below to explain key patterns. Most of the mean values were neutral (neither agree or disagree) (Table 5). Only a few items had a high mean value > 4: *"I want to have a say in my risk assessment and risk management"* (M: 4.62, SD: 0.78); *"I feel it is important that I am involved in my risk assessment and risk management"* (M: 4.68, SD: 0.73); and *"there needs to be better communication between professionals and service users about risk*" (M: 4.59, SD: 0.78). Service users’ attitudes towards friends and family supporting their involvement in RA and RM was ambivalent, as the negative and positive statements both received a mean value < 3. The statement *"my mental health team help me to be involved in my risk assessment and risk management"* also received a low mean value (M: 2.68, SD: 1.36). Interestingly, the first three items measuring participants’ involvement and awareness of their RA and RM content had a mean value < 3.

**Figure 2.**
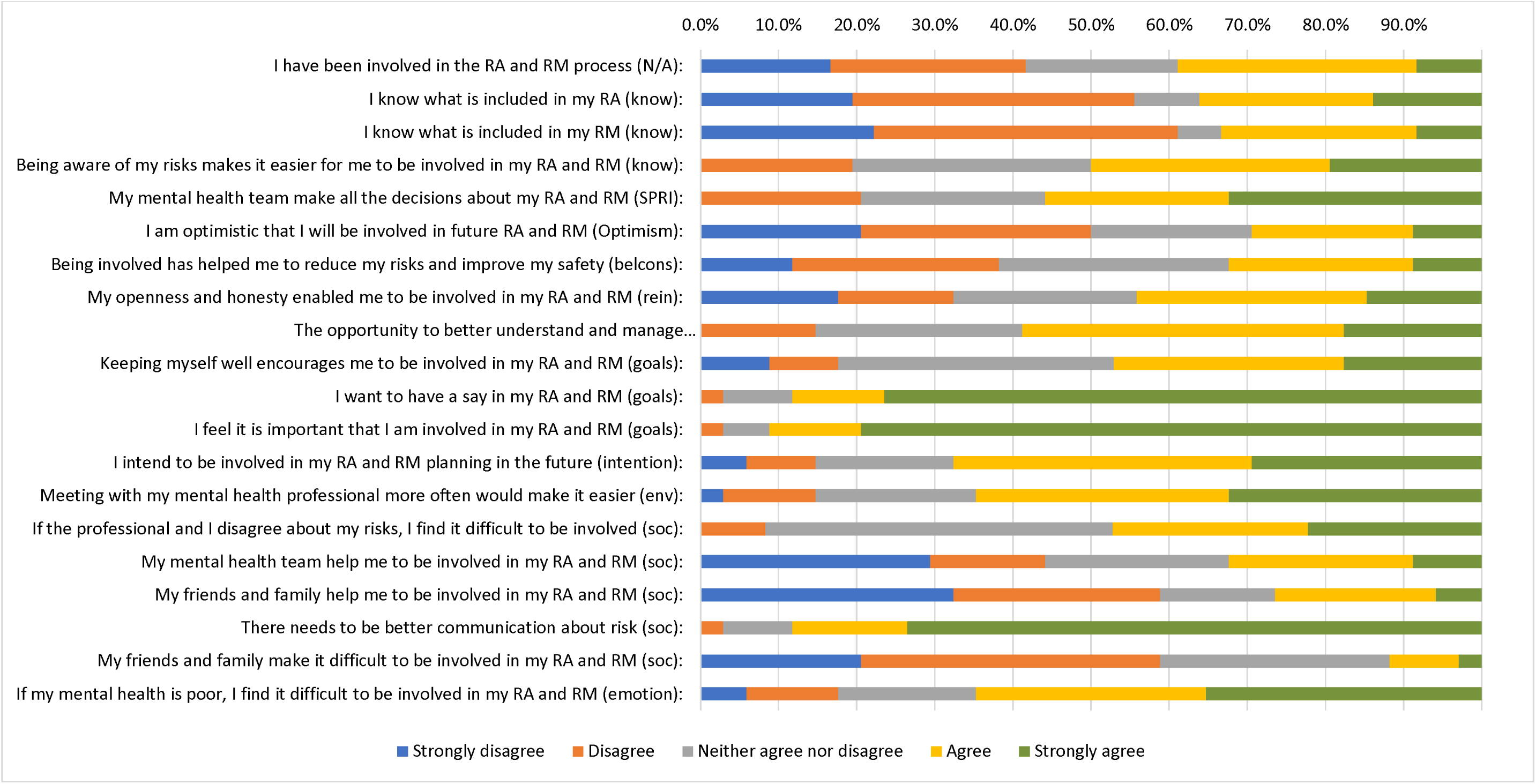
Responses to questions based on the lived experience belief statements (n = 36)

[INSERT Table 5. Mean and standard deviation for each item (lived-experience survey)]

### Mental Health Professionals’ Responses to the Theoretical Domains Framework Survey

#### Internal Consistency of Questionnaire Domains

Across the nine domains, Cronbach’s alpha values ranged from 0.56 (goals) to 0.80 (beliefs about consequences). Six domains met the threshold for acceptable internal consistency (α >0.7), while three fell below this level: Skills = 0.67, Goals = 0.56, and Emotions = 0.58) but their mean inter-item correlations value were within the optimal range: Skills = 0.29, Goals = 0.39, Emotions = 0.41. Further item-level analysis was conducted to explore whether internal consistency could be improved by removing specific items (See Additional file 1, Appendix F for further detail).

#### Domain level analysis

Figure 3 illustrates the responses to each of the 39-items of the TDF questionnaire. In reporting the frequencies below, the positive responses (strongly agree and agree) and negative responses (strongly disagree and disagree) are reported together. Mean scores for the domains were all ≥ 3:00, indicating generally favourable views of participants towards implementing shared decision-making in risk assessment and management. Goals and reinforcement had the highest domain mean values (See Additional file 1, Appendix G).

**Figure 3.**
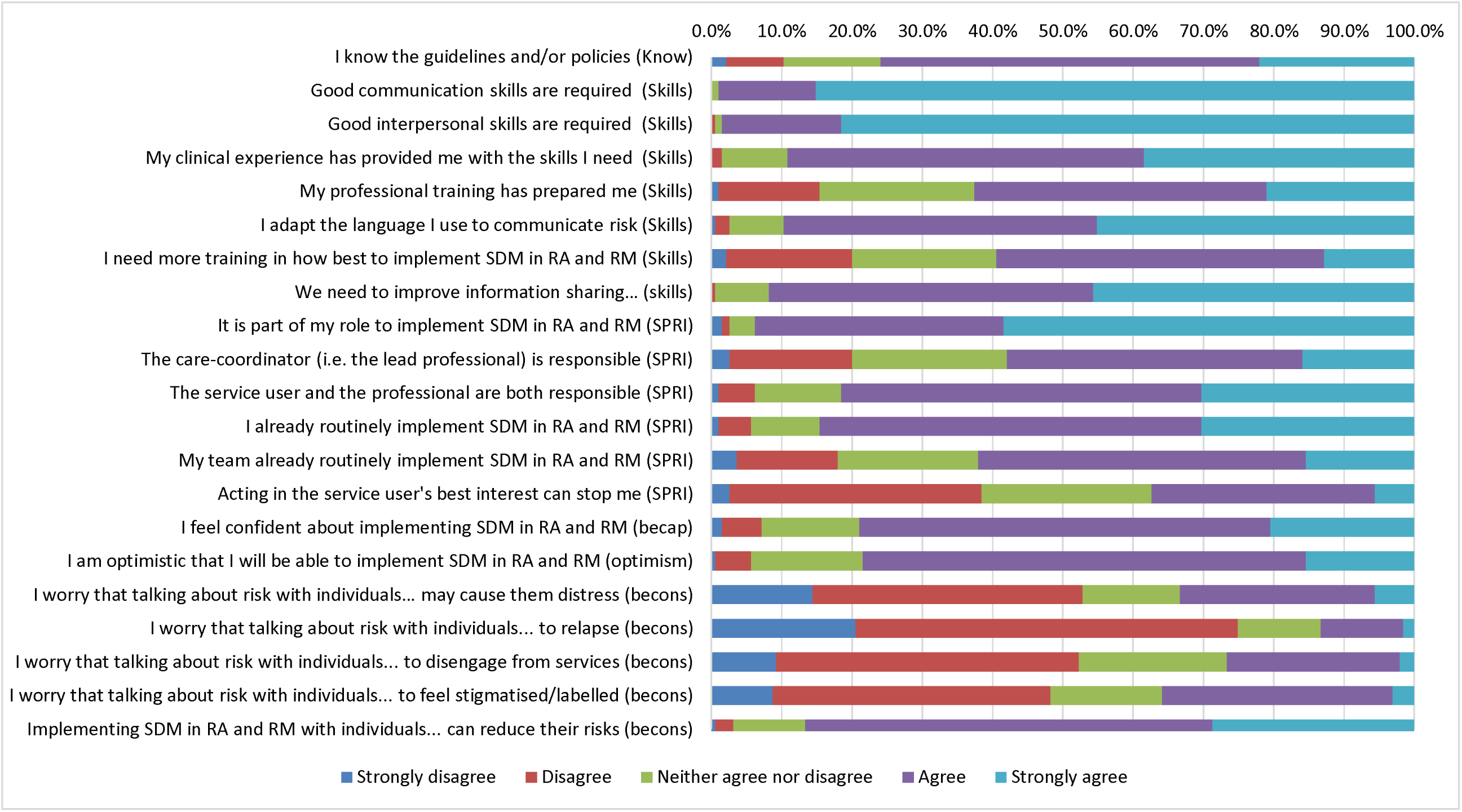
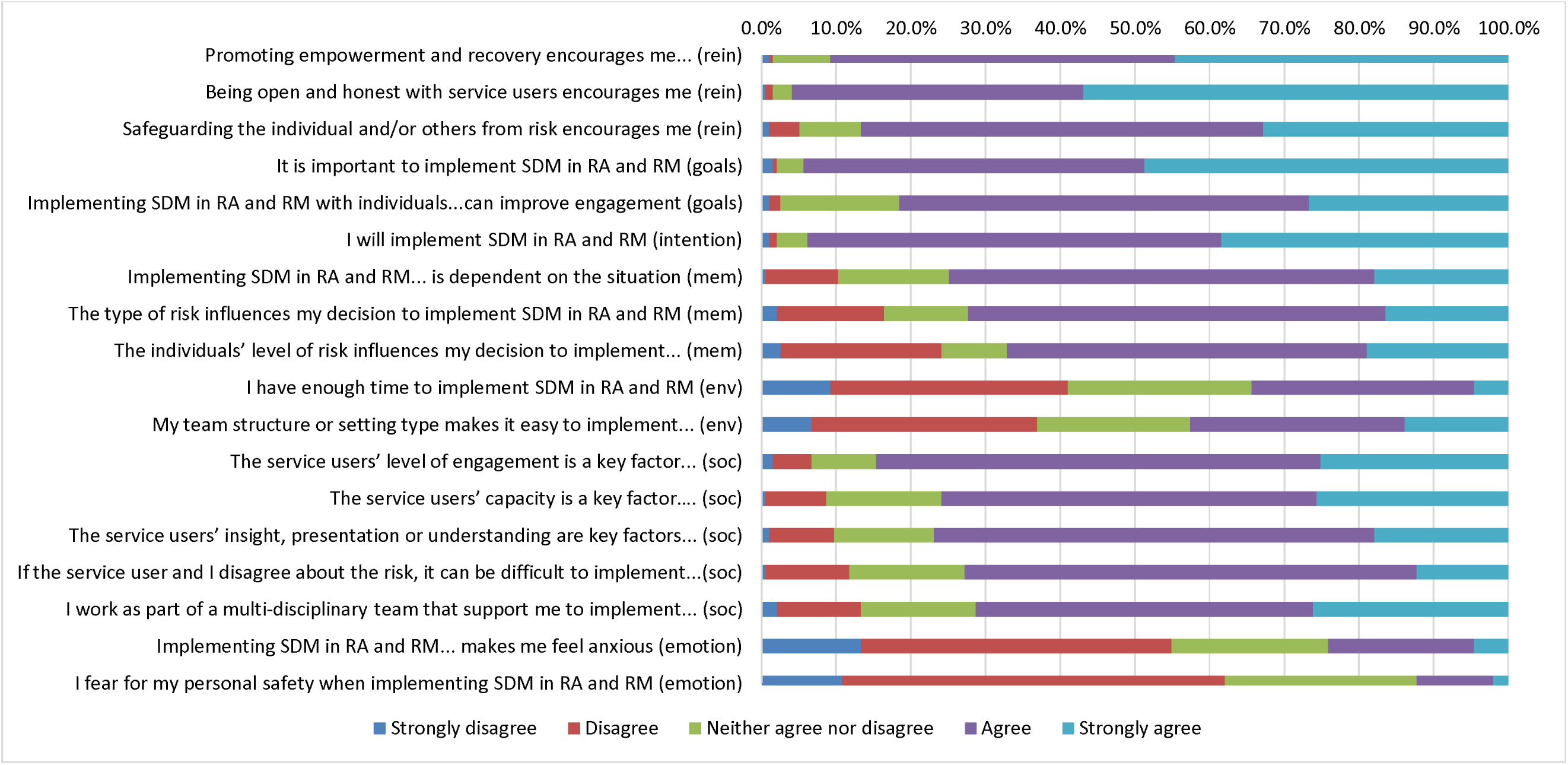
Mental Health Professionals Perceived Barriers and Enablers to SDM in RA and RM (n=195). ^1^SDM, Shared Decision-Making; RA, Risk Assessment; RM, Risk Management; SMI, severe mental illness. Know, knowledge; SPRI, social professional role and identify; becap, beliefs about capabilities; becons, beliefs about consequences; rein, reinforcement; mem, memory attention and decision making; env, environmental and resource factors; soc, social influences

#### Knowledge

A single item represented the domain of knowledge in the survey. The majority (75%) of those surveyed reported knowing the guidelines and/or policies that recommend SDM in RA and RM.

#### Skills

Responses were generally positive (strongly agree or agree). Almost all participants (98%) believed that good communication and interpersonal skills were required to implement SDM in RA and RM. There was also a high level of agreement (91.8%) towards the statement *"we need to improve information sharing between professionals about their experiences of implementing SDM in RA and RM"*. The majority of the items had a mean value > 4, indicating that skills were perceived as enablers to implementation (**Error! Reference source not found.**).

#### Beliefs about capabilities and optimism

Beliefs about capabilities and optimism were stand-alone domains containing one item each. Most participants were confident (79%) and optimistic (78.5%) that they would be able to implement SDM in RA and RM.

#### Social, professional role and identity

Almost all participants (93.9%) perceived it as part of their role to implement SDM in RA and RM, 81.6% also perceived it as a joint responsibility between themselves and the service user. A majority (84.7%) indicated that they were already implementing SDM in RA and RM. The lowest level of agreement (strongly agree and agree, 37.4%) was for the item *"acting in the service users’ best interest can stop me from implementing SDM in RA and RM"*.

#### Beliefs about consequences

Four out of five of the items in this domain were negatively worded statements. Three-quarters of the respondents disagreed (strongly disagree and disagree) that talking about risk could cause the service user to relapse. A lower level of disagreement was reported towards the statements *"I worry that talking about risk could: "…cause the service user distress or alarm"* (52.9%), *"…to disengage from services"* (52.3%) or *"…to feel stigmatised and/or labelled"* (48.2%). The item means for the above statements were < 3, indicating that these factors were generally not perceived as barriers to implementation. There was, however, a high level of agreement (strongly agree and agree, 86.6%) with the positive statement that *"Implementing SDM in RA and RM with individuals with SMI can reduce their risks"*.

#### Reinforcement

The domain of reinforcement received primarily positive responses (strongly agree and agree), with a high mean domain score of 4.32 (SD: 0.62). Participants believed that "promoting empowerment and recovery" (90.8%), ‘being open and honest’ (95.9%) and "safeguarding the individuals and/or others" (86.6%) encouraged them to implement SDM in RA and RM.

#### Goal and intention

There was a high level of agreement (94.3%) from respondents towards the statement *’It is important to implement SDM in RA and RM’. The* majority reported being willing (93.9%) to implement SDM in RA and RM.

#### Memory and decision making

Three-quarters of the participants perceived implementing SDM in RA and RM as situational dependent (74.8%) and considered the type of risk (72.3%) as an influencing factor. More than half agreed (strongly agree and agree, 67.2%) that the service users’ level of risk influences the implementation of SDM in RA and RM.

#### Environmental context and resources

More participants disagreed (41%) than agreed (34.3%) with the statement *’I have enough time to implement SDM in RA and RM’,* the mean value for this statement was < 3 indicating this as a potential barrier to implementation. Less than half (42.5%) of the participants also felt that their team structure or setting type made it easy for them to implement SDM in RA and RM.

#### Social influences

The majority of the items relating to social influences received positive responses (strongly agree and agree). For most participants (84.6%), being able to engage with service users was a key factor in implementing SDM in RA and RM. For three-quarters of respondents, the service user’s capacity (75.9%) and the service user’s insight, presentation or understanding (76%) were key factors in implementing SDM in RA and RM. Over two-thirds of the sample (71.3%) reported that their multidisciplinary team supported them to implement SDM, 72.8% believed it was difficult to implement SDM when they and the service user disagreed about risk.

#### Emotions

Over half of the participants disagreed (strongly disagree and disagree) that implementing SDM in RA and RM made them feel anxious (54.8%). The majority (62.1%) also disagreed with the statement that "I fear for my personal safety when implementing SDM in RA and RM". The latter negative statement’s mean value was < 3, suggesting that this was not perceived as a barrier to implementation (**Error!** Reference source not found.).

#### Principal components analysis

A principal component analysis (PCA) was conducted to reduce the 39 TDF items into a smaller set of components representing underlying factors. Four components were retained, representing the domains of: (1) motivation, (2) social influences and memory/decision-making, (3) beliefs about consequences, and (4) team, environment and training factors.

Responses to items of the four components are given in Table 6. Only statistically significant differences in component scores by demographic variables are reported below (see Additional file 1, Appendix H for full results).

#### Component 1: Motivation

The items collated and selected for interpretation related to the TDF domain’s goal, reinforcement, intention, social professional role and identity and optimism. Respondents generally held positive views, with a median overall score of 34 (IQR 32–37), range possible 8–40 (midpoint 24), with 40 representing the highest possible positive score. The data was assessed as not normally distributed. Kruskal-Wallis and Mann-Whitney U test was used to analyse for group differences. No statistically significant difference in the mean component score between the groups was evident, p>0.05.

#### Component 2: Social influences and memory, attention and decision-making

The items collated and selected for interpretation related to the TDF domain’s social influences and memory, attention and decision-making. Respondents generally held positive views, with a mean overall score of 19 (SD, 3.13), range possible 5–25 (midpoint 15), with 25 representing the highest possible positive score. Scores were normally distributed, parametric tests were used (ANOVA and t-test). No statistically significant difference between the means was found, p>0.05.

#### Component 3: Beliefs about consequences

Respondents generally scored beliefs about consequences highly, which in effect mean they disagreed with the statements as items were all reversed scored. The component had a mean overall score of 14 (SD, 3.49), range possible 4–20 (12 midpoint), with 20 representing the highest possible score. As the data were normally distributed, parametric tests were used (ANOVA and t-test).

A one-way analysis of variance revealed there was a statistically significant difference at the p < 0.05 level in component 3 scores for the three age groups: F (2, 192) = 5.08, p = 0.01. The partial eta-squared (η^2^ = .05) was of medium size. Post-hoc comparisons using the Tukey HSD test indicated that the mean score for the age group ≤36 (M = 12.58, SD = 3.56) was significantly different from that of the age group 47+ (M = 14.48, SD = 3.24). Age group 37-46 (M = 13.79, SD = 3.43) did not differ significantly from either age group ≤36 or 47+.

There was also a significant difference in component 3 scores between the three groups of years qualified in profession: F (2, 192) = 5.41, p = 0.01. The partial eta-squared (η^2^ = .05) was of medium size. Post-hoc comparisons using the Tukey HSD test indicated that the mean score for those qualified for ≤6 years (M = 12.82, SD = 3.32) was significantly different from those qualified for more than 16 years (M = 14.70, SD = 3.41).

#### Component 4: Team, environment and training factors

Respondents generally held neutral views, with a mean overall score of 13.4 (SD, 2.64), range possible 4–20 (midpoint 12), with 20 representing the highest possible positive score. As the data were normally distributed, parametric tests were used (ANOVA and t-test). A one-way analysis of variance revealed a statistically significant difference at the p ≤ 0.05 level in component 4 scores for the three groups of years qualified in profession: F (2, 192) = 3.10, p = 0.047. The partial eta-squared (η^2^ = .03) was of small size. Post-hoc comparisons using the Tukey HSD test indicated that the mean score for participants qualified for 7-15 years (M = 12.90, SD = 2.53) was significantly different from those qualified for more than 16 years (M = 14.05, SD = 2.73).

[INSERT Table 6. Components extracted from PCA, Cronbach alpha values and descriptive statistics]

## Discussion

The aim of this study was to ascertain whether the barriers and enablers to shared decision-making in risk assessment and management identified in our previous interview studies (13, 17) are important to many mental health professionals and service users.

Previous studies have found that service users are often not involved in the assessment and management of the risk component of their care (12, 34), or have limited awareness of the content of these documents (4). In our survey, two thirds (66.6%) of service user respondents reported not having received a copy of their risk assessment or management plan, despite most (68.8%) indicating that risk and safety had been discussed with them, and approximately half reporting involvement in these processes. This disparity between involvement and access to documentation raises important questions about the purpose and utility of risk assessment and management. Indeed, studies elsewhere indicate that professionals may engage service users in risk assessment for procedural reasons, such as information gathering (35-37). Shared decision-making, however, requires all parties to be informed and have an opportunity to contribute meaningfully to the decision-making processes (38). Risk plays an important role in mental health care, influencing outcomes such as access to services, autonomy and therapeutic risk-taking. Given the implications for the individual, the risk assessment and management process require openness and transparency.

Our findings also highlight the emotional and communicative challenges service users may encounter when engaging in discussions about risk. While participants valued being involved in the assessment and management of risk, many agreed that risk is a difficult and emotive topic to discuss with professionals. A majority indicated that communication about risk needs to improve. Previous research shows that service users may also struggle to raise safety concerns with professionals at times (39), due to factors such as fear of not being listened to, potential repercussions, and the impact of mental ill health. Relational care approaches to assessing and managing risk where professionals prioritise building trust and interpersonal relationships, may support service users in engaging more openly in discussions about risk (40).

Mental health professional participants also recognised the importance of effective communication, particularly the need to adapt the language used when discussing risk with service users (89% in agreement). The language of risk has been identified as a barrier to communication for both professionals and service users. In Langan and Lindow (41), for example, mental health professionals reported being cautious in the language they used when talking with service users about risk, particularly when discussing risk to others, as they were aware of the stigmatising impact it could have on the individual. In other studies, service users have reported not being familiar with the language of risk instead preferring to talk about their safety (42). The rationale for adapting language is justifiable, particularly when the intention is to communicate better with the service user or to support engagement. Our previous interview study (17), however, identified other reasons such as to avoid causing the service user distress or a lack of confidence in approaching sensitive topics. However, these motivations were not reflected in the current survey findings with most professionals reporting confidence in their ability to discuss risk with service users (79%, agree/strongly agree), and disagreeing with the belief about consequences survey items, including *‘I worry that talking about risk could cause the service user distress or alarm’* (52.9%).

User-led research also provides valuable insight into how risk and safety are perceived in mental health care. Faulkner (43) argues that the language of risk in mental health care is often pre-determined by professionals and does not reflect service users’ concerns. These themes have been reported elsewhere (37, 42), where professionals and service users conceptualised the term risk differently. Collaboratively identifying risk could broaden both stakeholders perception and also provide an opportunity to share concerns about risk that might otherwise be unknown (34). Faulkner (43) suggests that raising professionals’ awareness of the risks that service users themselves fear or experience may promote understanding of the importance of service users’ involvement in the decision-making process.

Nonetheless, professionals were generally positive about implementing shared decision making in risk assessment and management. However, a substantial proportion (41%) believed they did not have enough time to effectively implement SDM within these contexts. The finding is consistent with previous research, which has highlighted that professionals’ often lack time and opportunity to meaningfully collaborate with service users (44, 45). Also, working in pressured environments has been found to impact professional’s motivation to engage with service users (45) and increase coercive practices (46). Professionals, therefore, need support and time to build a relationship of trust with service users.

Principal component analysis of items measuring mental health professionals perceived barriers and enablers identified four components. The four components were labelled: ‘motivation’; ‘social influences and memory, attention and decision-making’; ‘beliefs about consequences’; and ‘team, environment and training factors’. Three of the components gained primarily positive scores. The scores for ‘team, environment and training factors’ were more negative. Those who had been qualified for longer in their profession scored higher in ‘team, environment and training factors’. This same group were also less influenced by their negative beliefs about consequences, such as causing distress, relapse, disengagement and stigma, than younger and less experienced professionals. In a study about professionals’ experiences of assessing and managing risk (47), it was believed that the skills needed to assess and manage risk were developed in practice over time, and the training received was deemed inadequate. Higgins, Doyle (3) also highlighted a need for risk assessment and safety planning guidance in education to enable professionals to work with risk within a collaborative recovery ethos. The present findings support the need for training in education to prepare younger and newly qualified professionals’ in implementing SDM and potentially reduce their negative beliefs about consequences. Also, less experienced professionals may benefit from ongoing clinical supervision and mentoring by more senior professionals.

### Strengths and limitations

A key strength of the study was the high completion rate for the mental health professional survey, which indicates interest in the topic and ease of completing the questionnaire. The survey was anonymous, and an incentive was offered to both groups. However, the recruitment rate for the service user survey was relatively low (n=48), and less than the estimated sample size needed for the planned statistical tests. Consequently, analysis of the data was limited to descriptive statistics. Recruitment difficulties in this population are well-documented. For example, the 2024 Community Mental Health Survey overall response rate was 20% (48). Factors such as survey length, sensitivity of the topic (e.g. risk), and previous findings that service users experience difficulty raising safety concerns (39) may have contributed to the low response rate in the present study. Nonetheless, this study is exploratory in nature and opens avenues for further research.

Another limitation of this study is the recruitment method. The survey was an open online survey that was posted primarily on social media. As millions of people visit social media websites, this method has the advantage of reaching a vast pool of participants and is considered appropriate for exploratory research (49). However, many people are still not engaged with social media, and even those who are may not have seen the advertisement to participate in the study. Second, controlling who takes an open online survey is impossible as the method relies on people to volunteer to the research process (49). Therefore, this method is prone to sampling biases, and the results obtained will have limited external validity (generalisability) (50).

The sample for service users also contained a higher proportion of individuals who identified as female (81.3%), and as White British (83.3%). This demographic pattern is consistent with findings from the 2024 Community Mental Health Survey (48), with females representing 62% of all respondents and most respondents identifying as white (87%). In addition, nearly a third of respondents in the current survey selected ‘other’ for their diagnosis and reported diagnoses including personality disorder, anxiety, depression, post-traumatic stress disorder and anorexia. As the survey items were developed from the key barriers and enablers identified in the interviews with people with SMI, the validity of the current findings would have been best if tested with people from the same population.

Finally, this study was limited by its inability to compare the experience of service users from different backgrounds. Previous research has shown that men from ethnic minority groups, particularly those from Black Caribbean or Black African backgrounds, are over-represented in mental health services (51), and disproportionately detained under the Mental Health Act 1983 (52). However, individuals from these groups are less likely to participate in mental health research compared to White counterparts (53). Barriers to participation cited in the literature include the stigma of mental illness, distrust of researchers and language barriers (53). To improve representation in future studies, researcher should consider recruiting directly from healthcare services using a probability sampling method to limit bias and increase representativeness. Additionally, strategies such as engaging with local community groups, and offering a translator may help increase participation among minority ethnic groups (53).

## Data Availability

All data produced in the present work are contained in the manuscript

