## Supplementary material for "Perceived Factors Influencing Shared Decision-Making in Mental Health Risk Assessment and Management: A Cross-Sectional Survey with Service Users and Professionals": Table 1

Table 1. Eligibility criteria for cross-sectional survey

| **Service users** | **Mental health professionals** |
| --- | --- |
| Aged 18 years or over | A qualified mental health professional (i.e., psychiatrist, psychologist, nurse, social worker or occupational therapist) |
| Receiving care and treatment from a community mental health service (i.e., providing care and support to people aged between 18 and 65) | Working within a community mental health team (i.e., providing care and support to people aged between 18 and 65) |
| Living with a severe mental illness (defined as psychosis, schizophrenia, bipolar disorder, severe depression) | Providing care to service users diagnosed with severe mental illness (defined as psychosis, schizophrenia, bipolar disorder, severe depression) |
|  | If their role involved assessing and managing risk with individuals with severe mental illness. |
