## Supplementary material for "Perceived Factors Influencing Shared Decision-Making in Mental Health Risk Assessment and Management: A Cross-Sectional Survey with Service Users and Professionals": Table 2

Table 2. Service user demographic and clinical characteristics (n=48)

| **Age in years**, mean (SD) | 38 (10.7) |
| --- | --- |
| **Gender**, n (%) |  |
| Female | 39 (81.3) |
| Male | 8 (16.7) |
| Prefer to describe | 1 (2.1) |
| **Ethnicity**, n (%) |  |
| White, British | 40 (83.3) |
| White, Irish | 1 (2.1) |
| White, other | 4 (8.3) |
| Mixed/Multiple ethnic, White and Asian | 1 (2.1) |
| Mixed/Multiple ethnic, other | 1 (2.1) |
| Asian, Pakistani | 1 (2.1) |
| **SMI diagnosis**, n, % (may have more than one diagnosis) |  |
| Schizophrenia | 4 (5.9) |
| Schizoaffective disorder | 4 (5.9) |
| Bipolar disorder | 16 (23.5) |
| Psychotic disorder | 9 (13.2) |
| Depression with psychotic features | 14 (20.6) |
| Other | 22 (30.9) |
| **Time in mental health services**, n (%) |  |
| Less than 1 year | 8 (16.7) |
| 1-3 years | 7 (14.6) |
| 4-6 years | 7 (14.6) |
| 7-9 years | 4 (8.3) |
| 10+ years | 22 (45.8) |
| **Type of mental health service**, n (%) |  |
| Early Intervention Service | 2 (4.2) |
| Any other Community Mental Health Service | 41 (85.4) |
| Both | 5 (10.4) |
| **Length of treatment from current service**, n (%) |  |
| Less than 1 year | 15 (31.3) |
| 1-3 years | 15 (31.3) |
| 4-6 years | 7 (14.6) |
| 10+ years | 11 (22.9) |
| **Relationship status**, n (%) |  |
| Single | 27 (56.3) |
| In a relationship | 9 (18.8) |
| Married | 11 (22.9) |
| Prefer not to say | 1 (2.1) |
| **Employment**, n (%) (multiple choice) |  |
| Full-Time employment | 9 (16.1) |
| Part-Time employment | 8 (14.3) |
| Education/Training | 4 (7.1) |
| Unemployed | 19 (32.1) |
| Voluntary work | 10 (16.1) |
| Other | 8 (14.3) |
| **Frequency of contact with family and friends**, n (%) |  |
| Daily | 25 (52.1) |
| Weekly | 10 (20.8) |
| Fortnightly | 6 (12.5) |
| Monthly | 2 (4.2) |
| Other | 5 (10.4) |
