## Supplementary material for "Perceived Factors Influencing Shared Decision-Making in Mental Health Risk Assessment and Management: A Cross-Sectional Survey with Service Users and Professionals": Table 3

Table 3. Mental health professional demographic characteristics (n=195)

| Age in years, mean (SD) | 42 (10.1) |
| --- | --- |
| Gender, n (%) |  |
| Female | 173 (88.7) |
| Male | 20 (10.3) |
| Prefer not to say | 2 (1) |
| Ethnicity, n (%) |  |
| White, British | 157 (80.5) |
| White, Irish | 7 (3.6) |
| Any other White | 10 (5.1) |
| Mixed/Multiple, White and Black | 2 (1.0) |
| Mixed/Multiple, White and Asian | 5 (2.6) |
| Any other Mixed | 1 (0.5) |
| Black, African | 5 (2.6) |
| Black, Caribbean | 2 (1.0) |
| Any other Black | 1 (0.5) |
| Asian, Indian | 2 (1.0) |
| Asian, Pakistani | 2 (1.0) |
| Arab | 1 (0.5) |
| Country of practice, n (%) |  |
| England | 152 (77.9) |
| Northern Ireland | 2 (1.0) |
| Scotland | 7 (3.6) |
| Wales | 16 (8.2) |
| Other | 18 (9.2) |
| Type of community service, n (%) |  |
| Early intervention service | 16 (8.2) |
| Any other community mental health | 168 (86.2) |
| Both | 11 (5.6) |
| Profession, n (%) |  |
| Mental Health Nurse | 62 (31.8) |
| Social Worker | 71 (36.4) |
| Occupational Therapist | 24 (12.3) |
| Psychologist | 15 (7.7) |
| Psychiatrist | 5 (2.6) |
| Other | 18 (9.2) |
| Years qualified in profession, n (%) |  |
| <1 year | 19 (9.7) |
| 1-3 years | 22 (11.3) |
| 4-6 years | 30 (15.4) |
| 7-9 years | 21 (10.8) |
| > 10 years | 103 (52.8) |
| Years working in current role, n (%) |  |
| <1 year | 29 (14.9) |
| 1-3 years | 91 (46.7) |
| 4-6 years | 32 (16.4) |
| 7-9 years | 11 (5.6) |
| > 10 years | 32 (16.4) |
| Are you a care-coordinator? n (%) |  |
| Yes | 98 (50.3) |
| No | 97 (49.7) |
