## Supplementary material for "Perceived Factors Influencing Shared Decision-Making in Mental Health Risk Assessment and Management: A Cross-Sectional Survey with Service Users and Professionals": Table 4

Table 4. Mean and standard deviation for each item (hypothetical)

| TDF Domain | Hypothetical belief statement | Mean | SD |
| --- | --- | --- | --- |
| **Knowledge** | *I am not aware of the risk assessment and risk management process: | 4.58 | 0.52 |
|  | *I do not know what is included in my risk assessment: | 4.75 | 0.45 |
|  | *I do not know what is included in my risk management: | 4.92 | 0.29 |
| **Skills** | *Talking about my risks and safety would be difficult: | 3.42 | 1.17 |
|  | Talking about my risks and safety would be easy: | 2.67 | 1.16 |
| **Social professional role and identity** | My risks are low, so I do not need to be involved in my risk assessment and risk management: | 2.42 | 1.00 |
|  | *My risks are high, so I do not need to be involved in my risk assessment and risk management: | 1.70 | 0.48 |
| **Beliefs about capabilities** | I would feel confident being involved in my risk assessment and risk management: | 4.42 | 0.67 |
| **Beliefs about consequences** | I would have been able to better understand and manage my risks if I was involved: | 4.17 | 0.72 |
|  | Being involved in my risk assessment and/or risk management may have helped me to reduce my risks and improve my safety: | 4.42 | 0.67 |
|  | I think having more involvement in my risk assessment and risk management would help me keep well: | 4.17 | 0.72 |
|  | Being involved in my risk assessment and risk management would keep me safe: | 4.00 | 0.82 |
| **Reinforcement** | My openness and honesty would have enabled me to be involved in my risk assessment and risk management: | 4.33 | 0.65 |
|  | I feel it is important that I am involved in my risk assessment and risk management: | 4.58 | 0.67 |
| **Goals** | I feel it is important that I have a say in my risk assessment and risk management: | 4.58 | 0.67 |
| **Intention** | I would have been willing to be involved in my risk assessment and risk management: | 4.50 | 0.67 |
| **Social Influences** | I think my friends and family would support me to be involved in my risk assessment and risk management: | 3.80 | 1.55 |
|  | *We need better communication between service users and professionals about risk: | 5.00 | 0.00 |
| **Emotion** | *Talking about my risks would make me feel distressed and/or upset: | 3.67 | 1.07 |
|  | I am more likely to be involved in my risk assessment and risk management when my mental health is poor: | 3.00 | 1.63 |
|  | I believe I have not been involved in my risk assessment and risk management because my mental health has been good: | 2.60 | 0.97 |

SD, Standard deviation, strongly disagree = 1 to strongly agree = 5, *negative statement, reversed scored
