## Supplementary material for "Perceived Factors Influencing Shared Decision-Making in Mental Health Risk Assessment and Management: A Cross-Sectional Survey with Service Users and Professionals": Table 5

Table 5. Mean and standard deviation for each item (lived-experience survey)

| **TDF Domain** | **Lived-experience belief statement** | Mean | SD |
| --- | --- | --- | --- |
| N/A | I have been involved in the risk assessment and risk management process: | 2.89 | 1.26 |
| **Knowledge** | I know what is included in my risk assessment: | 2.75 | 1.38 |
|  | I know what is included in my risk management: | 2.58 | 1.32 |
|  | Being aware of my risks makes it easier for me to be involved in my risk assessment and risk management: | 3.50 | 1.03 |
| **Social Professional role and identity** | *My mental health team make all the decisions about my risk assessment and risk management: | 3.68 | 1.15 |
| **Optimism** | I am optimistic that I will be involved in future risk assessment and risk management: | 2.68 | 1.27 |
| **Beliefs about consequences** | Being involved in my risk assessment and/or risk management has helped me to reduce my risks and improve my safety: | 2.91 | 1.16 |
| **Reinforcement** | My openness and honesty enabled me to be involved in my risk assessment and risk management: | 3.09 | 1.33 |
|  | The opportunity to better understand and manage my risks encourages me to be involved in my risk assessment and risk management: | 3.62 | 0.95 |
| **Goals** | Keeping myself well encourages me to be involved in my risk assessment and risk management: | 3.38 | 1.16 |
|  | I want to have a say in my risk assessment and risk management: | 4.62 | 0.78 |
|  | I feel it is important that I am involved in my risk assessment and risk management: | 4.68 | 0.73 |
| **Intention** | I intend to be involved in my risk assessment and risk management planning in the future: | 3.76 | 1.16 |
| **Environmental context and resources** | Meeting with my mental health professional more often would make it easier to be involved in my risk assessment and risk management: | 3.79 | 1.12 |
| **Social Influences** | *If the professional and I disagree about my risks, I find it difficult to be involved in my risk assessment and risk management: | 3.61 | 0.93 |
|  | My mental health team help me to be involved in my risk assessment and risk management: | 2.68 | 1.36 |
|  | My friends and family help me to be involved in my risk assessment and risk management: | 2.41 | 1.31 |
|  | *There needs to be better communication between service users and professionals about risk: | 4.59 | 0.78 |
|  | *My friends and family make it difficult to be involved in my risk assessment and risk management: | 2.35 | 1.01 |
| **Emotion** | *If my mental health is poor, I find it difficult to be involved in my risk assessment and risk management: | 3.76 | 1.23 |

SD, Standard deviation, strongly disagree = 1 to strongly agree = 5, *negative statement, reversed scored
