## Supplementary material for "Perceived Factors Influencing Shared Decision-Making in Mental Health Risk Assessment and Management: A Cross-Sectional Survey with Service Users and Professionals": Table 6

Table 6. Components extracted from PCA, Cronbach alpha values and descriptive statistics

| Statement | Responses n (%) | | | | |
| --- | --- | --- | --- | --- | --- |
|  | Strongly disagree | Disagree | Neither agree nor disagree | Agree | Strongly agree |
| **Component 1 – Motivation (8 items)** | | | | | |
| It is important to implement SDM in RA and RM | 3 (1.5) | 1 (0.5) | 7 (3.6) | 89 (45.6) | 95 (48.7) |
| Being open and honest with service users encourages me to implement SDM in RA and RM | 1 (0.5) | 2 (1.0) | 5 (2.6) | 76 (39.0) | 111 (56.9) |
| I will implement SDM in RA and RM | 2 (1.0) | 2 (1.0) | 8 (4.1) | 108 (55.4) | 75 (38.5) |
| Promoting empowerment and recovery encourages me to implement SDM in RA and RM | 2 (1.0) | 1 (0.5) | 15 (7.7) | 90 (46.2) | 87 (44.6) |
| It is part of my role to implement SDM in RA and RM | 3 (1.5) | 2 (1.0) | 7 (3.6) | 69 (35.4) | 114 (58.5) |
| Safeguarding the individual and/or others from risk encourages me to implement SDM in RA/RM | 2 (1.0) | 8 (4.1) | 16 (8.2) | 105 (53.8) | 64 (32.8) |
| I am optimistic that I will be able to implement SDM in RA and RM | 1 (0.5) | 10 (5.1) | 31 (15.9) | 123 (63.1) | 30 (15.4) |
| I already routinely implement SDM in RA and RM | 2 (1.0) | 9 (4.6) | 19 (9.7) | 106 (54.4) | 59 (30.3) |
| Cronbach statistics, sum of allocating 1 (strongly disagree) to 5 (strongly agree)  Cronbach alpha 0.89  Range possible 8–40, with 40 representing highest positive score  Mid-point 24  Median 34  IQR 32-37 |  |  |  |  |  |
| **Component 2 – Social influences and Memory, attention and decision making (5 items)** | | | | | |
| The service users' insight, presentation or understanding are key factors in whether I can implement SDM in RA and RM | 2 (1.0) | 17 (8.7) | 26 (13.3) | 115 (59.0) | 35 (17.9) |
| The service users' capacity is a key factor in whether I can implement SDM in RA and RM | 1 (0.5) | 16 (8.2) | 30 (15.4) | 98 (50.3) | 50 (25.6) |
| Implementing SDM in RA and RM with an individual with SMI is dependent on the situation | 1(0.5) | 19 (9.7) | 29 (14.9) | 111 (56.9) | 35 (17.9) |
| The individuals' level of risk influences my decision to implement SDM in RA and RM | 5 (2.6) | 42 (21.5) | 17 (8.7) | 94 (48.2) | 37 (19.0) |
| The service users' level of engagement is a key factor in whether I can implement SDM in RA & RM | 3 (1.5) | 10 (5.1) | 17 (8.7) | 116 (59.5) | 49 (25.1) |
| Cronbach statistics, sum of allocating 1 (strongly disagree) to 5 (strongly agree)  Cronbach alpha 0.75  Range possible 5–25, with 25 representing highest positive score  Mean 19.2  SD 3.13 |  |  |  |  |  |
| **Component 3 – Belief about consequences (4 items)** | | | | | |
| *I worry that talking about risk with individuals with SMI may cause them distress or alarm | 28 (14.4) | 75 (38.5) | 27 (13.8) | 54 (27.7) | 11 (5.6) |
| *I worry that talking about risk with individuals with SMI may cause them to relapse | 40 (20.5) | 106 (54.4) | 23 (11.8) | 23 (11.8) | 3 (1.5) |
| *I worry that talking about risk with individuals with SMI may cause them to disengage from services | 18 (9.2) | 84 (43.1) | 41 (21.0) | 48 (24.6) | 4 (2.1) |
| *I worry that talking about risk with individuals with SMI may cause them to feel stigmatised and/or labelled | 17 (8.7) | 77 (39.5) | 31 (15.9) | 64 (32.8) | 6 (3.1) |
| Cronbach statistics, sum of allocating 1 (strongly disagree) to 5 (strongly agree)  Cronbach alpha 0.84  Range possible 4–20, with 20 representing highest positive score  Mean 13.5  SD 3.49 |  |  |  |  |  |
| **Component 4 – Team, environment and training factors (4 items)** | | | | | |
| My team structure or setting type makes it easy to implement SDM in RA and RM | 13 (6.7) | 59 (30.3) | 40 (20.5) | 56 (28.7) | 27 (13.8) |
| My team already routinely implement SDM in RA and RM | 7 (3.6) | 28 (14.4) | 39 (20.0) | 91 (46.7) | 30 (15.4) |
| *I need more training in how best to implement SDM in RA and RM with individuals with SMI | 4 (2.1) | 35 (17.9) | 40 (20.5) | 91 (46.7) | 25 (12.8) |
| I have enough time to implement SDM in RA and RM | 18 (9.2) | 62 (31.8) | 48 (24.6) | 58 (29.7) | 9 (4.6) |
| Cronbach statistics, sum of allocating 1 (strongly disagree) to 5 (strongly agree)  Cronbach alpha 0.73  Range possible 4–20, with 20 representing highest positive score  Mean 13.4  SD 2.64 |  |  |  |  |  |

*negative statement, reversed scored; SDM, Shared Decision Making; RA, Risk Assessment; RM, Risk Management; SD, Standard Deviation
